# Efficacy of Probucol on cognitive function in Alzheimer’s disease: Study protocol for a double-blind, placebo-controlled, randomised phase II trial (PIA Study)

**DOI:** 10.1101/2021.11.20.21266372

**Authors:** Virginie Lam, Roger Clarnette, Roslyn Francis, Michael Bynevelt, Gerald F Watts, Leon Flicker, Carolyn Orr, Poh Loh, Nicola T Lautenschlager, Christopher M Reid, Jonathan K Foster, Satvinder Dhaliwal, Suzanne Robinson, Emily Corti, Mauro Vaccarezza, Ben Horgan, Ryusuke Takechi, John C.L Mamo

**Affiliations:** Curtin Health Innovation Research Institute, Faculty of Health Sciences, Curtin University, Bentley, Australia; School of Population Health, Faculty of Health Sciences, Curtin University, Bentley, Australia; Australian Alzheimer’s Research Foundation, University of Western Australia, Nedlands, Australia; School of Medicine, University of Western Australia, Crawley, Australia; Department of Nuclear Medicine, Sir Charles Gairdner Hospital, Nedlands, Australia; Neurological Intervention and Imaging Service of Western Australia, Sir Charles Gairdner Hospital, Nedlands, Australia; Cardiometabolic Service, Department of Cardiology and Internal Medicine, Royal Perth Hospital, Perth, Australia; WA Centre for Health and Ageing, University of Western Australia, Crawley, Australia; Royal Perth Hospital, Perth, Australia; Academic Unit for Psychiatry of Old Age, Department of Psychiatry, The University of Melbourne, Melbourne, Australia; North Western Mental Health, Royal Melbourne Hospital, Parkville, Victoria, Australia; Division of Psychiatry and WA Centre for Health and Ageing, University of Western Australia, Perth, Australia; Synapse Neuropsychology, Perth, Australia; Faculty of Health Sciences, Curtin University, Bentley, Australia; School of Paediatrics and Child Health, Faculty of Health and Medical Science, University of Western Australia, Crawley, Australia; Department of Radiation Oncology, Sir Charles Gairdner Hospital, Nedlands, Australia; Duke-NUS Medical School, National University of Singapore, Singapore; Institute for Research in Molecular Medicine (INFORMM), Universiti Sains Malaysia, Pulau Pinang, Malaysia; Curtin Medical School, Faculty of Health Sciences, Curtin University, Bentley, Australia

## Abstract

**Introduction:** Preclinical, clinical and epidemiological studies support the hypothesis that aberrant systemic metabolism of amyloid-beta (Aβ) in the peripheral circulation is causally related to the development of Alzheimer’s disease (AD). Specifically, recent studies suggest that increased plasma concentrations of lipoprotein-Aβ compromises the brain microvasculature, resulting in extravasation and retention of the lipoprotein-Aβ moiety. The latter results in an inflammatory response and neurodegeneration ensues.

Probucol, a historic cholesterol-lowering drug, has been shown in murine models to suppress lipoprotein-Aβ secretion, concomitant with maintaining blood-brain-barrier function and suppressing neurovascular inflammation. Probucol has also been shown to protect cognitive function in dietary-induced amyloidogenic mice.

This protocol details the Probucol in Alzheimer’s Study (PIA-study), a double-blind, randomised, placebo-controlled drug intervention trial investigating if Probucol attenuates cognitive decline in patients with mild-to-moderate AD.

**Objectives:** The primary objective of the 104-week study is to assess whether Probucol supports cognitive function and delays brain atrophy in AD patients. A secondary objective is to determine whether Probucol treatment will reduce cerebral amyloid burden.

**Methods & Analysis:** The study is a phase II single-site, randomised, placebo-controlled, double-blind clinical trial assessing the efficacy of Probucol in AD. A total of 300 participants with mild-to-moderate AD will be recruited and randomised 1:1 (active: placebo). Cognitive function, regional volumetric changes in brain and cerebral amyloid load will be evaluated via the cognitive subscale test, AD assessment scales (ADAS-Cog), magnetic resonance imaging (MRI) and positron emission tomography (PET) scans, respectively, after a 104-week intervention.

**Ethics & Dissemination:** The study has been approved by the Bellberry Limited Human Research Ethics Committee (Approval number: HREC2019-11-1063; Version 4, 6^th^ October 2021). The investigator group will disseminate study findings through peer-reviewed publications, key conferences and local stakeholder events.

**Trial registration:** This trial has been registered with the Australian New Zealand Clinical Trial Registry (ACTRN12621000726853).

**ARTICLE SUMMARY:** *Strengths and limitations of this study:*

- This is the first-in-human (FIH) randomised double-blind placebo-controlled study to assess the efficacy of Probucol in delaying cognitive decline in individuals with mild cognitive impairment (MCI) and mild-to-moderate dementia due to Alzheimer’s disease (AD).
- The 24-month intervention study will be the first to investigate whether treatment with Probucol will stabilise structural/functional changes in brain and if cerebral amyloid load will decrease in individuals with AD, following treatment with Probucol.
- Probucol is clinically used to treat cardiovascular disease with well-characterised efficacy and safety profiles, thus reducing risk of the study, and if applicable, accelerate clinical translation of the study findings.

## INTRODUCTION

### Background and rationale

Alzheimer’s disease (AD) is a neurodegenerative disorder affecting approximately 50 million people worldwide. Extracellular deposition of amyloid beta (Aβ) is a hallmark pathological feature of AD featuring prominently within the hippocampal formation and entorhinal cortex. Amyloidosis is positively associated with cognitive decline in AD and targeting amyloidosis presently is a therapeutic priority^1^. Recently, the US Food and Drug Administration (FDA) Federal Drug Agency approved Aducanumab, a treatment which decreases amyloid plaque burden in some patients with AD^2, 3^.

Microvascular disturbances are the first pathological feature of AD that may include microbleeds; hypo-perfusion and or blood-brain barrier dysfunction with changes in the extracellular matrices associated with astrogliosis^4, 5^. Contemporary treatments for AD include cholinesterase inhibitors such as Galantamine, Rivastigmine, or Donepezil, to support synaptic activity, or Memantine to regulate glutamate^6^. Remarkably, of approximately 400 clinical trials in AD, to our knowledge, none to date have targeted the brain microvasculature, or peripheral metabolism of Aβ to reduce risk for, or progression of AD.

Several recent epidemiological studies show that systemic measures of Aβ in blood positively correlate with cerebral amyloid burden and cognitive decline in AD^7, 8^. A causal association is suggested by findings that plasma Aβ isoforms discriminate with a high degree of accuracy in individuals who go on to develop AD, decades before onset of disease^8^. However, presently the mechanism(s) by which peripheral Aβ metabolism might exacerbate AD risk are not well understood. Insight into how blood Aβ increases risk for AD comes from findings that in humans, greater than 90% of blood Aβ_1-40_ and 97% of the particularly pro-amyloidogenic Aβ_1-42_ is associated with plasma lipoproteins^9^, principally the triglyceride-rich lipoproteins (TRL) of hepatically derived very-low-density-lipoproteins (VLDL) and of postprandial chylomicrons^10^. To directly address the hypothesis of a lipoprotein-Aβ/capillary axis for AD, mice were engineered to synthesise human Aß restricted exclusively to the liver. Lam *et al*. reported marked neurodegeneration concomitant with capillary dysfunction, parenchymal extravasation of lipoprotein-Aß, and neurovascular inflammation^11^. The liver-specific amyloid mice also showed impaired performance in the passive avoidance test, suggesting impairment in hippocampal-dependent learning. Collectively, these findings provide a strong rationale to consider interventions that target and modulate peripheral metabolism of lipoprotein-Aβ to mitigate AD risk.

Probucol is an historic and safe cholesterol lowering drug, clinically used in Japan since 1985, with potent anti-inflammatory and anti-oxidant properties^12-14^. Probucol was also shown to profoundly attenuate dietary induced synthesis and secretion of lipoprotein-Aβ^15, 16^ concomitant with cerebral capillary integrity sparing^17^. In a dietary-induced diabetic murine model, Probucol was also found to support hippocampal-dependent memory recall^18^. The pleiotropic properties of Probucol and significant clinical use experience justifies considering repurposing Probucol to test efficacy in supporting cognitive function in patients with AD.

We describe the Probucol in Alzheimer’s-study (PIA-Study). The study is a phase II placebo-controlled, double-blind clinical trial assessing the efficacy of Probucol in AD. Key outcome measures include cognitive function, regional volumetric changes in brain and cerebral amyloid load.

## OBJECTIVES

The primary objective of this study is to evaluate the efficacy of Probucol (Lorelco™) on cognitive performance in AD patients over a 104-week treatment period. The secondary objectives are [1] to evaluate regional volumetric changes and cerebral amyloid abundance in the brain of AD patients treated with Probucol (Lorelco™) over a 104-week treatment period, [2] to evaluate improvement or maintenance of quality-of-life parameters in patients with AD, and [3] to assess the safety and tolerability of Probucol (Lorelco™) in patients with AD.

## METHODS AND ANALYSIS

### Trial design

This is a single-site, phase II, randomised, double blind, placebo-controlled parallel group study in adults with mild-to-moderate AD. The study will assess the efficacy, safety and tolerability of the treatment of AD individuals with Lorelco™. Participants, study doctors and researchers will be blinded to allocation of the study medication. The maximum study duration is 112 weeks (2 years, 8 weeks), with a treatment period of 104 weeks. There is a 4-week screening phase to ensure all participants have measurable mild or moderate AD and to ensure eligibility in the study. There is also a 4-week follow-up after the end of the treatment period. We aim to recruit 300 participants for this study. Eligible participants will be randomised in a 1:1 (active: placebo) ratio using permuted block randomisation. Eligible participants will be randomised to a unique participant code, which will be assigned to a participant number. Participant numbers will be randomised to either Probucol (Lorelco™) or placebo manufactured by Oxford Compounding. The trial coordinator will randomly assign the participant’s screening number to a unique participant code at Week 1, Day 1. Participants will be dosed as follows; week 1 and 2: 1 x placebo taken in the morning, with food; week 3: 1 × 250 mg Lorelco™ (or matching placebo) taken in the morning, with food; and week 4 – 104: 1 × 250 mg Lorelco™ (or matching placebo) taken in the morning and in the evening, with food. An overview of the study design is shown in **Figure 1** and the overall schedule of the trial is illustrated in **Table 3**.

**Figure 1.**
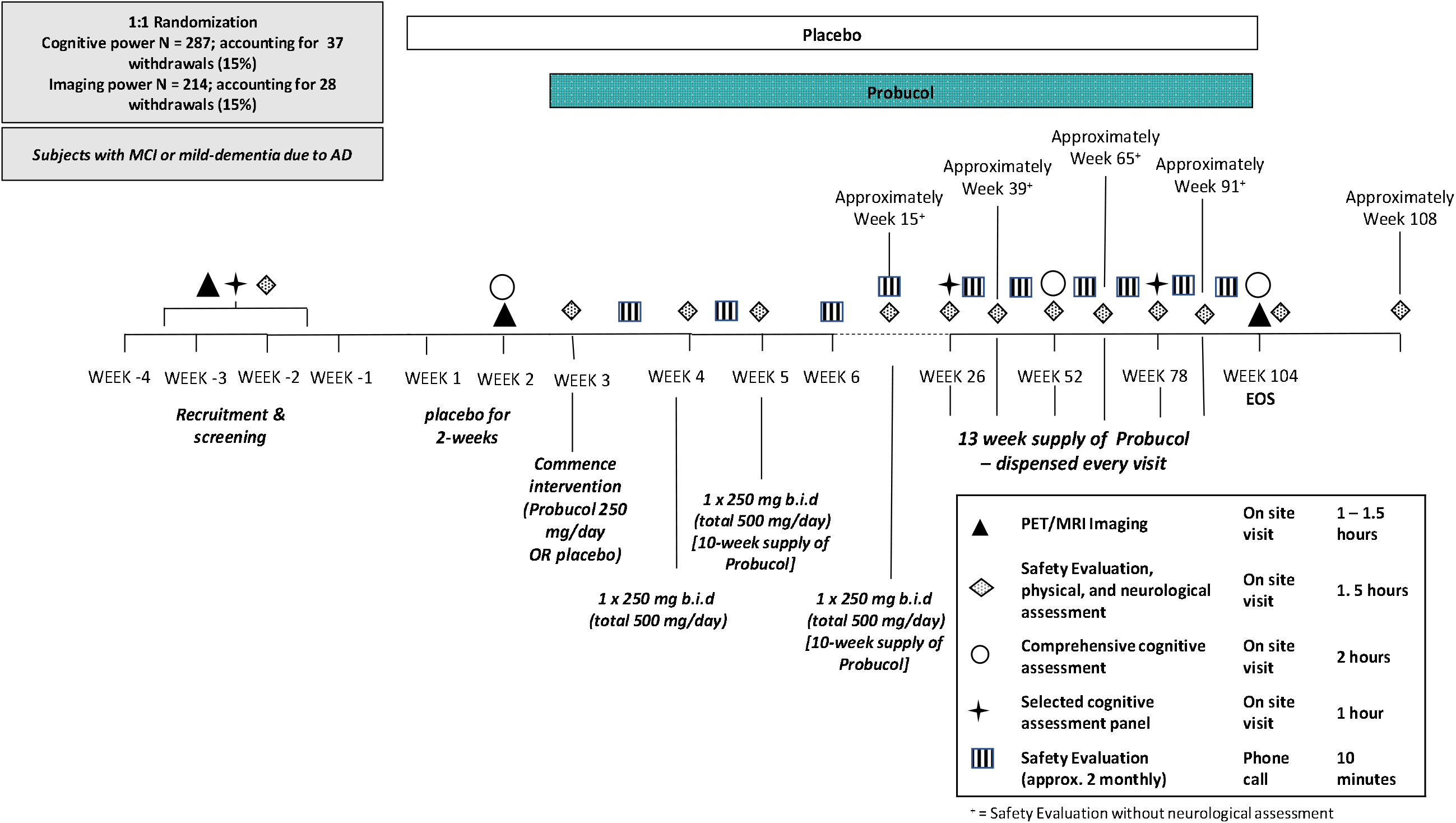
Overview of PIA trial study design

### Study setting and recruitment

This study will be based in Australia. All assessments and blood collection will be completed at the Australian Alzheimer’s Research Foundation (AARF) based at Hollywood Specialist Centre, Nedlands, Western Australia. Positron Emission Tomography (PET) imaging will be completed at Sir Charles Gairdner Hospital, Nedlands, Western Australia. Magnetic Resonance Imaging (MRI) will be completed at Envision Medical Imaging, Perth, Western Australia. To reach the targeted sample size, participants will be identified and recruited from the investigators’ private and public patient clinics; referrals from GP or other specialists; phone calls to site via word of mouth; the Alzheimer’s Association Research Foundation website; or via database review where participants who have previously given permission for the site to contact them to inform them with new studies. Participants will also be recruited through advertising and editorials on social, print, radio and/or television media; correspondence and promotions via not-for-profit organisations and research partners; website and newsletter content.

### Eligibility criteria

Individuals will be eligible for the study if they meet all of the inclusion criteria and do not satisfy any of the exclusion criteria listed in **Table 1**.

**Table 1.**
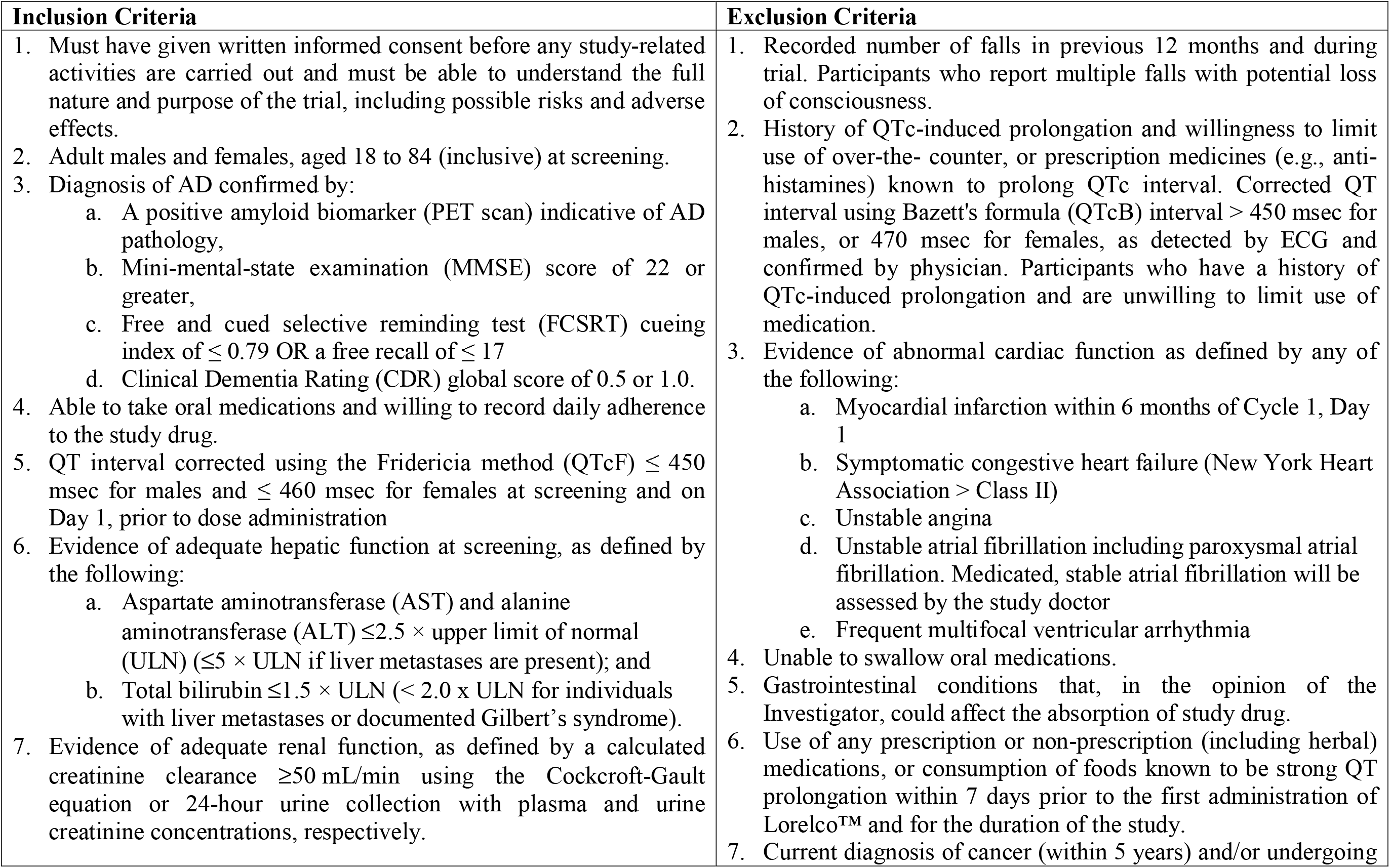
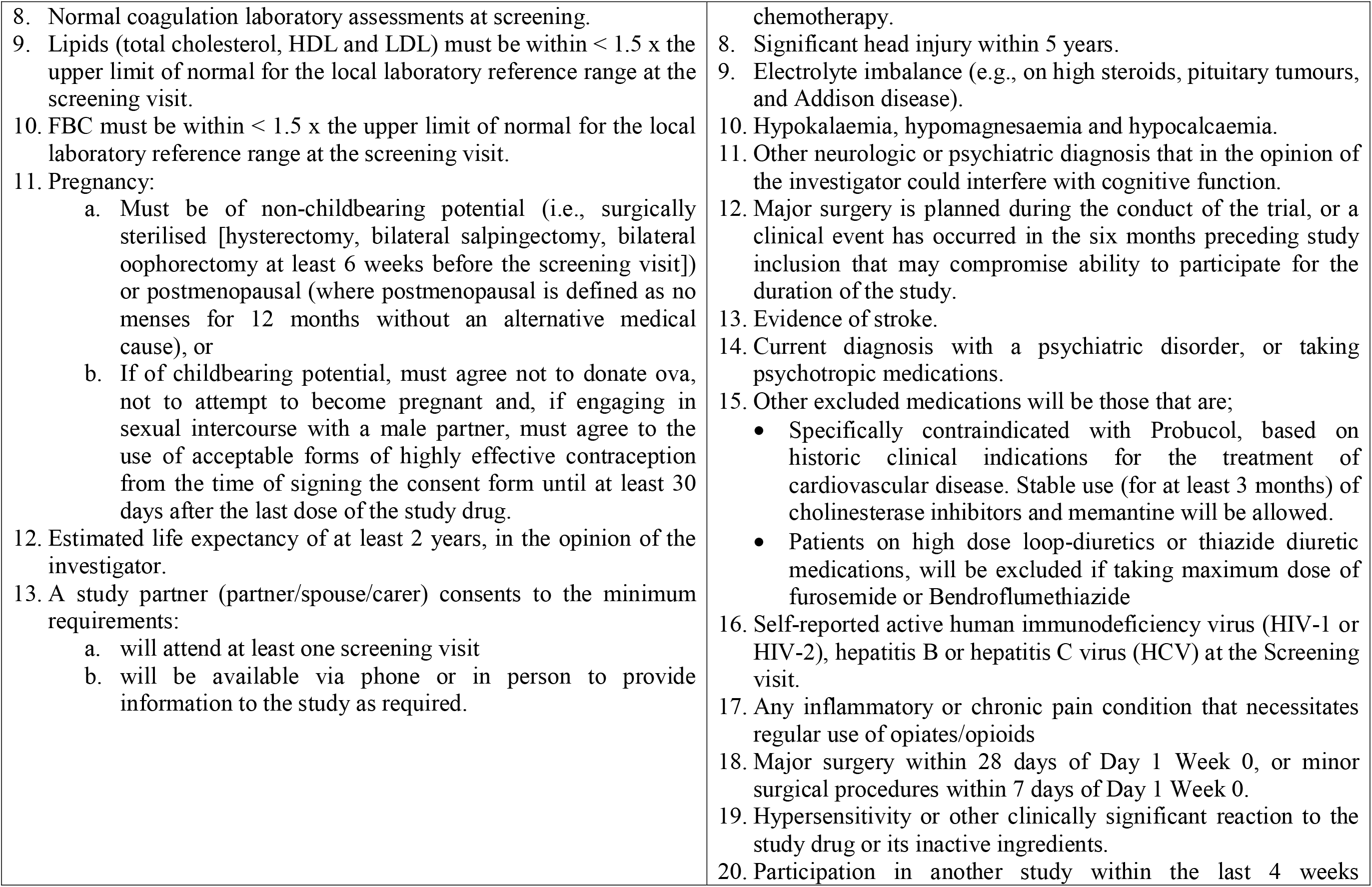
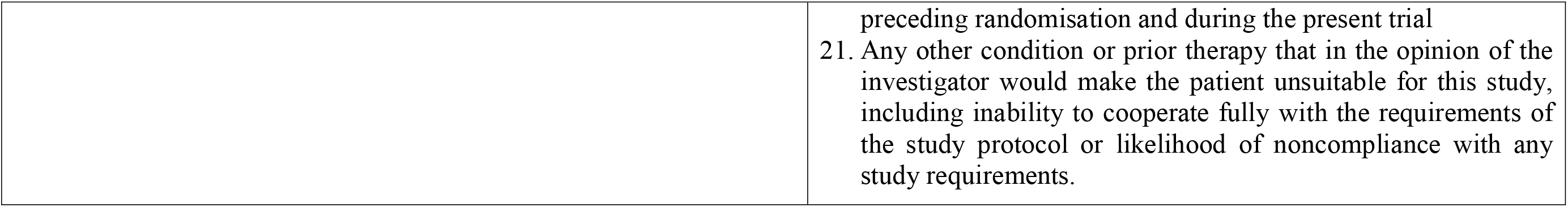
PIA trial inclusion and exclusion criteria

## Intervention

### Intervention description

During the initial recruitment and screening phase, patients will be screened against the exclusion criteria to ensure suitability for participation. During the screening phase, participants will undergo a short cognitive screening assessment, safety assessment, and complete a PET scan to determine cerebral amyloid load. Study medication will commence with a single dose escalation design with all participants receiving initially for two-weeks 1 X placebo consumed with food, after which baseline measures for cognitive performance, structural and functional brain MRI scans will be completed. Thereafter, (week 3) patients receive for one week 1 X Lorelco™ (250 mg) capsule (or matching placebo), taken with food. Commencing week 4, patients receive Lorelco™ 250 mg b.i.d, consumed with food. Study medication will be dispensed at every study visit. Safety evaluations throughout the study will comprise physical/neurological examinations, ECG, vital sign measurements, standard laboratory tests, and monitoring for adverse events at weeks 3, 4, 5, 15, 26, 39, 52, 65, 78, 91, 104, and 108. At weeks 3, 4, 6, 20, 29, 47, 55, 73, and 81, the study coordinator will contact the patient or caregiver by phone record to determine any adverse events.

### Supply of study drug

Probucol used in this study will be commercially available tablets, Lorelco™, produced by Aventis Pharmaceuticals and wholesaled by Otsuka Pharmaceutical Co., Ltd. Lorelco™ tablets (250 mg) will be over-encapsulated inside an opaque capsule shell and backfilled with microcrystalline cellulose. Individual doses of Lorelco™ will be dispensed by the site pharmacy. Matching placebo opaque capsules with no active ingredients and a filler of microcrystalline cellulose will be compounded by Oxford Compounding.

### Safety

Protocol violations should not lead to treatment discontinuation unless they pose a significant risk to participant safety. Trial stopping criteria, dose stopping rules and individual dosage adjustments are indicated in **Table 2**.

**Table 2.** Criteria PIA trial stopping criteria and intervention dosage adjustments

|  |
| --- |
| ***Trial stopping criteria*** |
| ○ Substantial deviations from the approved protocol<br>○ Adverse-effects of unexpected type, severity, or frequency are encountered<br>○ As the trial progresses, the continuation of the trial would disadvantage some of the participants as determined by the investigator group, trial monitors or the data safety and monitoring committee (DSMB).<br>*All participants will take part in a safety evaluation 4 weeks following discontinuation. |
| ***Dose stopping criteria and individual dosage adjustments*** |
| Dosing may be stopped or modified if suspected adverse drug reactions, changes in vital signs, ECGs, or clinical laboratory results are observed, and these changes pose a significant health risk. Of particular note for dose discontinuation are the following criteria;<br>○ Corrected QT/QTc interval is in excess of 500 ms, or if it increases > 50 ms compared to baseline (baseline will be calculated as the mean of two time points, screening and baseline day 1).<br>○ Occurrence of QT/QTc interval prolongation in association with symptoms of arrhythmia<br>○ Subsequent prescription of a drug with potential QTc prolongation (Antimalarial; Macrolides; Quinolones; Triazole antifungals; Anti-arrhythmic; Antiemetic; Anti-depressant; Antipsychotics)<br>○ Evidence of liver dysfunction, or primary biliary cirrhosis<br>○ If for any reason the investigator team determines that continued participation in the trial is not in the participant's best interest. |

### Adverse events

The investigators will report any serious adverse events (AEs) occurring during the clinical trial, independent of direct causal relationship with the treatment, within 24 hours. Unblinding will be permissible in the event information is required to ensure the participants safety in case of an AE. The study doctor will provide Oxford Compounding with the unique participant code to unblind the participant. All AEs will be reviewed by the independent DSMB, specifically appointed for the trial.

### Participant Withdrawal

If a participant decides to withdraw from the project, participants are asked to notify a member of the research team. If a participant withdraws consent during the research project, the study doctor and relevant project team members will not collect additional personal information from the participant. However, personal information already collected will be retained to ensure that the results of the research project can be measured properly and to comply with law. Participants will be made aware that data collected by the Sponsor up to the time the participant withdraws will form part of the research project results.

### Adherence

The two-week placebo period will serve to monitor participant adherence to the treatment schedule. Participants will also be required to return the study treatment webster packs at every clinic visit. In the event adherence falls below 80%, participants will be re-trained in the administration of the study medication. If adherence continues to be below 80% the participant will be withdrawn from the study.

### Concomitant care

This study allows for ‘usual clinical care’. Some medications or treatments may not be permitted during participation in this study. The study doctor will collect information about concomitant medication use during every study visit and safety screening. Participants will not be permitted to take part in other studies/investigational treatments for AD or other health conditions while taking part in this study.

### Study Procedures

For the exact timing of each procedure, please refer to the Schedule of Assessments (SoA; **Table 3**).

**Table 3.**
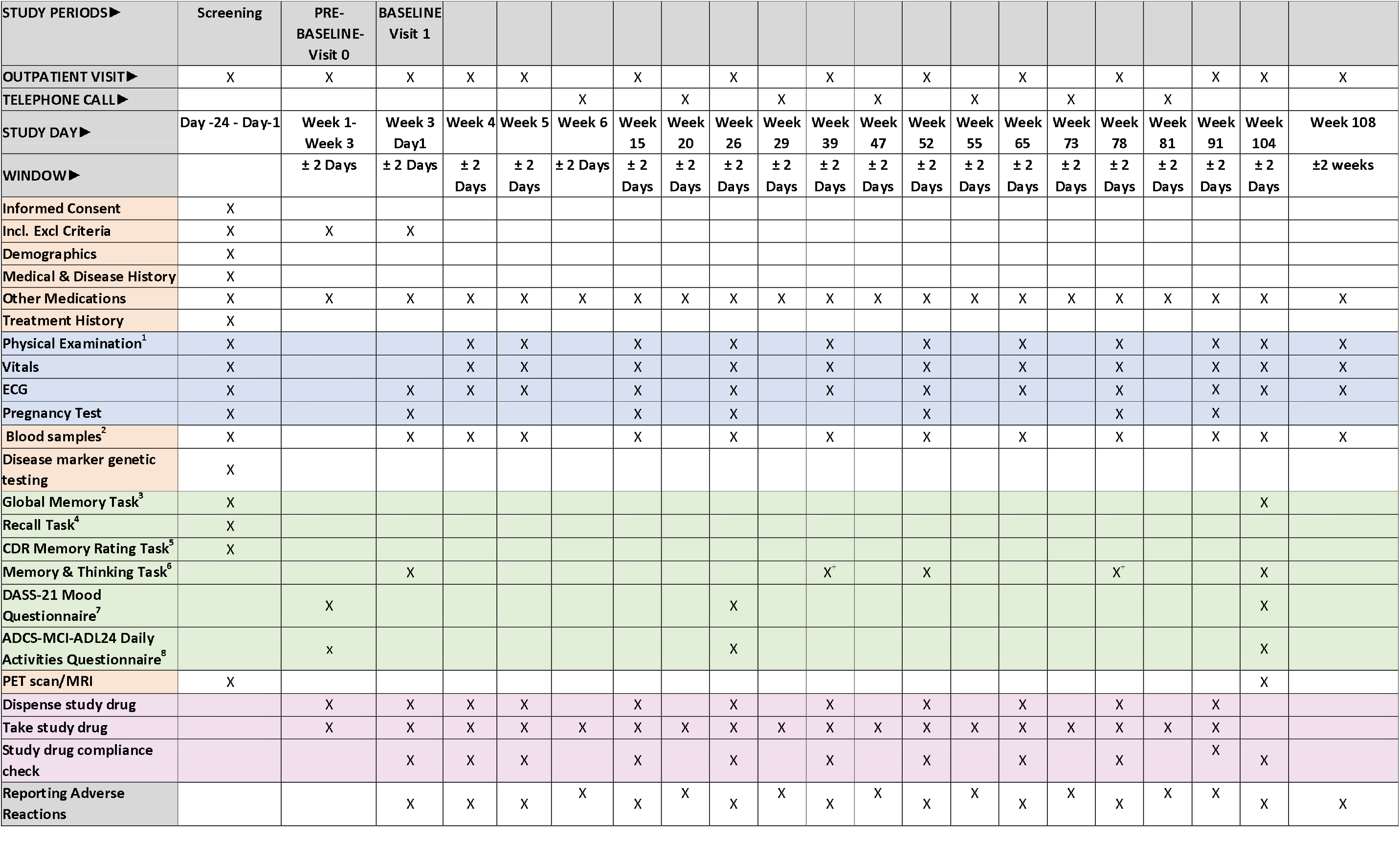
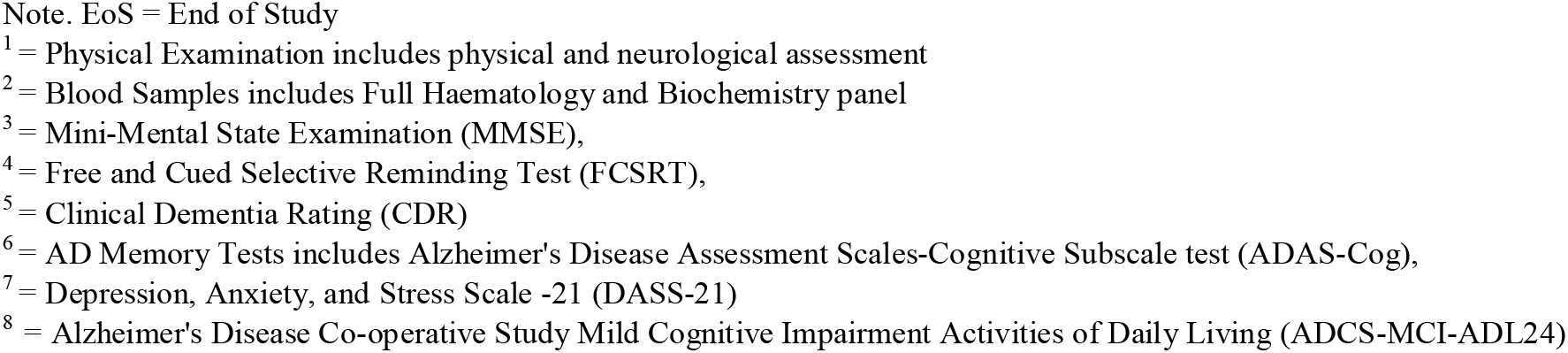
Overall Schedule of Assessments (SoA) for the PIA trial

### Screening Assessments

#### Informed Consent

The Information and Informed Consent Form (ICF) will be provided to patients at Screening and signed consent must be provided prior to any study procedures being performed.

#### Medical History

A full medical history will be obtained at screening, including a detailed neurologic history, other medical and surgical history, medication history and drug allergies. Demographic data including gender, ethnicity, and race will be recorded.

#### Height and Weight

Body height (centimetres) and weight (kilograms) will be measured, and body mass index (BMI) will be calculated.

#### Pregnancy Test

Female patients (women of child-bearing potential (WOCBP) only) will complete a urine chorionic gonadotropin (hCG) pregnancy test at screening, baseline, and again at EoS visit.

### Safety and Tolerability Assessments

Safety will be determined by evaluating physical and neurological examinations, vital signs, clinical laboratory parameters, 12-lead ECGs and AEs. Abnormal vital signs assessments, clinical laboratory safety tests, ECGs and physical examinations that are judged by the PI as clinically significant will be recorded as AEs or SAEs. The timing of all safety assessments is presented in **Table 3**.

#### Vital Signs

Vital signs assessments will include systolic and diastolic blood pressure, heart rate (HR), respiratory rate (RR) and body temperature. Patients should be resting in a supine position for at least 5 minutes prior to and during vital signs measurements.

#### Clinical Laboratory Safety Tests

Fasted blood samples (minimum 8 hour fast) will be collected by venepuncture at screening, baseline, weeks 3, 4, 5, 15, 26, 39, 52, 65, 78, 91 and 104 weeks (an estimated 13 mL of blood will be collected per visit and a total of 160 mL will be collected over the 2-year study).

Blood samples for haematology, serum biochemistry (including liver function tests) will be collected at selected time points throughout the study (see **Table 3**). Test results will be monitored for potential AEs, including gastrointestinal bleeding (haemoglobin) and rhabdomyolysis (plasma CK). Apolipoprotein E genotype will also be determined.

#### Electrocardiogram

Twelve-lead ECGs will be assessed (including but not limited to the measurements of ventricular HR, PR interval, RR interval, QRS duration, QT interval and QTcF). Screening and prior to first dose, triplicate 12-lead ECGs (collected within 5 minutes with each reading separated by at least 1 minute) will be taken to establish eligibility at baseline. Triplicate ECGs will also be recorded at the EoS visit. The average value for the triplicate will be utilised for assessing QTcF inclusion criteria. All other ECGs will be single readings. ECG normal ranges are as follows: PR interval: 120 msec – 220 msec (inclusive); QRS duration: < 120 msec; QTcF ≤450 msec (males); QTcF ≤460 msec (females); HR 45-100 beats/min (inclusive)

#### Physical Examination

A full physical examination will be performed at screening and at the EoS visit. The full physical examination will include, at a minimum, assessment of the following systems: skin, head, ears, eyes, nose and throat, lymph nodes, heart, chest, abdomen and extremities, and a neurological examination (assessment of speech, cranial nerves, peripheral nerves, motor power, deep tendon reflexes, sensation, coordination and gait) and any other focussed assessments suggested by the presence of specific symptoms. All other scheduled assessments will be symptom-directed.

### Cognitive Screening Assessments

#### Free and Cued Selective Reminding Test (FCSRT)^19^

The FCSRT assessment will be performed at screening only for eligibility determination. A cueing index of ≤ 0.79 is required for study entry. The FCSRT is a cued recall test that utilises a controlled encoding technique to ascertain that impairment in recall and cueing are due to a memory deficit rather than a failure of encoding.

#### Clinical dementia rating scale (CDR)^20^

The CDR provides two scores, a global score (GS) and a sum of boxes (SOB). The GS distinguishes a participant’s level of impairment into the following categories: 0 (normal); 0.5 (questionable dementia; 1 (mild dementia); 2 (moderate dementia) and 3 (severe dementia). The SOB is scored from 0 – 18 with higher scores indicating a greater level of impairment. The scale covers six domains: memory, orientation, judgment and problem solving, community affairs, home and hobbies and personal care.

#### Mini Mental State Examination (MMSE)^21^

The MMSE is a brief, widely used 30-item assessment of global cognition examining orientation, registration, calculation, recall, attention and language. The spelling of WORLD backwards will not be used in this protocol. Participants who score <22 at screening will be ineligible for study entry. MMSE will be assessed at screening and EoS.

All cognitive assessments will be performed by an independent assessor who will be blinded to treatment allocation.

### Outcome measures

#### Primary Outcome measures

The primary outcome measure will be the change in the Alzheimer’s Disease Assessment Scales-Cognitive Subscale test (ADAS-Cog)^22^. The ADAS-Cog is the most widely used test to measure cognition in RCT’s for AD. The ADAS-Cog consists of the following tasks: Word Recall Task; Following Commands; Constructional Praxis; Delayed Word Recall; Naming Objects and Fingers; Ideational Praxis; Orientation; Word Recognition; Spoken Language; Comprehension and Word Finding Difficulty. The full suite of ADAS-Cog will be assessed at baseline, 26, 52, 78 and 104 weeks.

An additional outcome measure will be assessment of brain morphometry and volume determined via magnetic resonance imaging (MRI) at baseline (pre-intervention) and EoS. The MRI protocol will include the acquisition of 3 sets of data [1] Volumetric isotropic T1 scan (6.5 min). This will allow voxel wise segmentation and volumetric analyses (e.g., grey matter volume) to assess volume changes in characteristic locations which can yield diagnostic accuracy on approximately 90%^23^. Mesial temporal lobe (hippocampus and entorhinal cortex via Scheltens grading), global cortical atrophy (Pasquier scale) and parietal atrophy (Koedam score) as well as inferior lateral ventricle size will be assessed. Brain volume indices indicate that patients with AD have accelerated rates of brain volume loss of up to 4.5% per year compared to normal controls (1%). [2] 3D FLAIR scan (3.5min). This will demonstrate the small vessel ischaemic lesion load which will be scored according to the Fazekas method^24^. [3] Susceptibility weighted imaging (SWI) – a means of measuring micro bleed load and indicator of amyloid angiopathy and for the purposes of Quantitative Susceptibility Mapping (QSM) (∼7 mins)^24^. This method quantitates regional brain iron content which is altered in AD compared with normal controls. Mesial temporal, basal ganglia, cingulate, cortical region of interest comparisons will be performed at baseline and at treatment completion. Visible micro bleeds on the SWI imaging will be graded using the Brain Observer Micro-Bleed Scale (BOMBS).

#### Secondary Outcomes

Quality of life will be assessed as a secondary outcome measure via the Alzheimer’s Disease Co-operative Study Mild Cognitive Impairment Activities of Daily Living (ADCS-MCI-ADL24)^25^. The ADCS-MCI-ADL24 is a study partner 24-item questionnaire evaluating perceived difficulties with functioning in several activities of daily living across a variety of domains. The Depression Anxiety Stress Scale (DASS-21), a self-report 21-item will be utilised to determine levels of depression, anxiety and stress^26^. Participants are read a statement and asked to rate each statement on a four-point scale as to how much it relates to them. DASS-21 will be administered at day 1/week 1, week 26 and EoS.

Cerebral amyloid load will be assessed as an additional imaging outcome measure. Brain amyloid imaging will be done with amyloid tracer positron emission tomography (PET) scans at baseline and at EoS. Dynamic and static PET imaging will be acquired. Analysis will include [1] visual assessment of amyloid load [2] quantitative assessment of amyloid burden, including standard uptake value ratio (SUVr) and, [3] dynamic imaging for blood perfusion measures.

## Statistics

### Estimated sample size and power

Estimated sample sizes are calculated for the two primary outcome measurements: ADAS-Cog and grey matter atrophy (hippocampal). In order to ensure that the study has sufficient power to detect differences in both of the primary outcomes, the sample size chosen is the maximum of that calculated for each primary outcome.

The primary analysis is an intention-to-treat analysis and will include all randomized participants. Data will be analysed using both Generalised Estimating Equations (GEE) and Bayesian Analysis. The analysis of primary endpoints will use linear mixed-effects models, with random slopes and intercepts. For the ADAS-Cog, using mixed model analysis published estimates from the ADNI cohort^27^ suggest a sample size of 125 AD participants per trial arm (total N = 250) will be required for power at 0.8 to detect a drug effect of 25% over two years and assuming a decline from baseline of 1.10 standardised units on the composite (SD change = 0.83). For the MRI markers, Ledig *et al*. reported the sample sizes required for a 25% intervention reduction over two years based on 322 patients with AD (with 117 followed for 24 months) and a reduction of 10.2% (6.2) for hippocampus^28^. Sample size calculations based on hippocampal volume suggest that 93 participants per treatment arm are required (total N = 186). Assuming a 20% attrition rate, a sample of 314 individuals will be recruited for the cognitive study, and 233 individuals will be randomly chosen for the imaging study.

### Statistical Analysis

#### Outcomes

The primary analysis is an intention-to-treat analysis of all randomized participants. Data will be analysed using both GEE and Bayesian Analysis. The analysis of primary endpoints will use linear mixed-effects models, with random slopes and intercepts. Analysis of all primary and secondary endpoints contrasting Probucol and placebo, after adjusting for covariates, will use mixed effects-regression with ‘random’ intercepts and slopes (as has been used for power calculations). Mean differences and associated 95% confidence intervals will be presented for the ‘fixed’ effect of Probucol treatment. No formal interim analyses are planned at this time.

#### Additional analyses

Further analysis investigating the relationship between change scores (post-minus pre-intervention scores) for the primary ADAS-Cog with MRI volumes (total grey matter, hippocampus, and medial temporal lobe volumes) and specific blood biomarkers (e.g.: plasma lipoprotein-Aβ) will be considered using Pearson (or Spearman where appropriate) correlation analysis. For all other correlations between recorded variables that lack an a priori hypothesis, control of statistical errors will be carried out using Holm-Sidak corrections for multiple comparisons.

If Probucol treatment is successful, a directed acyclic graph Bayesian network analysis will be carried out a posteriori on variables identified to be significant predictors of either grey matter arrest or neuropsychological performance to better elucidate mechanisms of the effect of Probucol. Greedy equivalence search will be used to identify statistical conditional dependencies between variables and directionality will be estimated using the linear, non-Gaussian, acyclic causal models (LiNGAM) approach^29, 30^. Goodness of fit will be estimated using a χ2 test contrasting the identified model against a saturated model.

In addition to Bayesian analyses, the traditional General Linear Model analysis will also be utilised to compare Probucol to placebo, after adjusting for covariates. The Generalized Estimating Equations method, which extends the generalized linear model to allow for analysis of repeated measurements or other correlated observations, will also be utilised. Missing data on the ADAS-Cog will not require data imputation. The scoring methodology for the ADAS-Cog as proposed by Verma *et al*. will be utilised as it estimates cognitive impairment using the set of items answered by the patients^22^. Any additional missing data will be identified using Missing Values Analysis and will be replaced using multiple imputation where appropriate. Mean difference and associated 95% confidence intervals will be presented. Data will be analysed using Stata Version 16.

### Data and safety monitoring

#### Data management

Data will be collected by study delegated personnel on paper source maintained in a participant study binder in secure facilities at Alzheimer’s Association Research Foundation (AARF). Identifiable data will be stored securely and kept in a locked cabinet with access restricted to the investigator team, site and monitoring personnel. All other data will be de-identified to sure confidentiality of participant data. Data will be stored electronically on password protected web-enabled clinical trial data electronic management system (REDCap) located in an ISO27001 compliant facility at Curtin University. Clinical records collected at recruitment will be kept in a locked cabinet in a locked office at the site and will collectively be housed in secure facilities at the AARF. Participants’ study information will not be released outside of the study without the written permission of the participant. All data will be securely archived as per the Sponsor’s data policy for a minimum of 25 years.

#### Trial monitoring and formal committees

The trial monitoring committee (TMC), comprised of the principal investigator, key trial staff including the trial manager, a nurse representative and a consumer representative, are responsible for trial setup, ongoing management and promotion of the trial. The trial steering committee (TSC), comprising the investigator team including geriatricians, cardiologists, neuroradiologists, nuclear medicine physicians, neuropsychologists, a biostatistician, clinical biochemists, consumer and community representatives, will provide overall supervision of the study and are responsible for interpretation and dissemination of results.

AARF employs independent data auditors who will monitor and audit compliance of data entry/management, legislation, regulations, guidelines and codes of practice, at quarterly intervals. Findings from each audit will be discussed with the study coordinator and thereafter with the investigator team to ensure any action items are addressed promptly and appropriately.

An independent data safety and monitoring committee (DSMB) will oversee the safety aspects of the study. The DSMB consists of members with expertise in clinical pharmacology, biostatistics, clinical trial design, and clinical cardiology. Members of the DSMB will not be investigators of the study nor will they have any conflict of interest with the investigators. The committee will meet periodically to advise the TSC on the progress of efficacy and safety data as it accumulates throughout the course of the study. The TSC and DSMB will provide independent oversight of the study.

### Ethics and dissemination

#### Ethics Approval

The PIA study has been approved by Bellberry Ltd Human Research Ethics Committee (HREC2019-11-1063). This trial has been registered with the Australian New Zealand Clinical Trial Registry (ACTRN12621000726853) and is registered in accord with the World Health Organization Trial Registration Data Set. The Universal Trial Number is U1111-1259-0486. Where applicable, approved protocol amendments will be communicated to the trial personnel and relevant committees.

### Informed consent and withdrawal from the study

Participants will be in the mild stages of dementia and therefore are expected to be able to provide informed consent. An investigator or research staff delegate will explain all study procedures and possible risks to the participant. The participant and their nominated study partner will have an opportunity to have all questions answered and thereafter will sign and date the ICF, indicating willingness to participate in the study. Participants will be informed prior to consent that they can withdraw at any time without their care being affected in any way.

### Dissemination

Participants will be updated about the progress and results of the study via presentations or newsletters from the investigator group. The results will be disseminated via peer-reviewed publications, key conferences and local stakeholder events.

### Trial Status

This study is in the process of recruiting participants and expected to complete in 2026.

### Patient and public involvement

The study was developed in consultation with a consumer advocate representing the Consumer and Community Involvement Program (https://cciprogram.org/). The trial will be overseen by the trial steering committee (TSC), including patient and public members.

## STUDY SPONSOR

Curtin University

Kent Street, Bentley, 6102

Western Australia

+61 8 9266 9266

## STUDY SPONSOR & FUNDER RESPONSIBILITIES

Curtin University, the National Health and Medical Research Council, and MSWA do not have any responsibility relating to study design; collection, management, analysis, and interpretation of data; writing of the report; and the decision to submit the report for publication.

## Data Availability

All data produced in the present study are available upon reasonable request to the authors.

## AUTHOR CONTRIBUTIONS

V.L, R.C, R.F, M.B, S.S.D, R.T and J.C.LM conceived the study concept. V.L, R.C, R.F, M.B, G.F.W, L.F, S.S.D, E.C, R.T and J.C.LM designed the trial protocol. L.F, C.O, P.L, N.T.L, C.M.R, J.K.F, S.R, M.V and B.H commented on the methods and contributed to the development of the study. V.L, E.C and J.C.L.M drafted the manuscript. All authors revised the manuscript and approved the final version.

## FUNDING STATEMENT

The PIA-trial is funded by the National Health and Medical Research Council Medical Research Future Fund for Neurological Disorders (MRF1201204), the Multiple Sclerosis Society of Western Australia (MSWA) and the McCusker Charitable Foundation.

## COMPETING INTERESTS

None declared.

### APPENDICIES

#### Appendix A

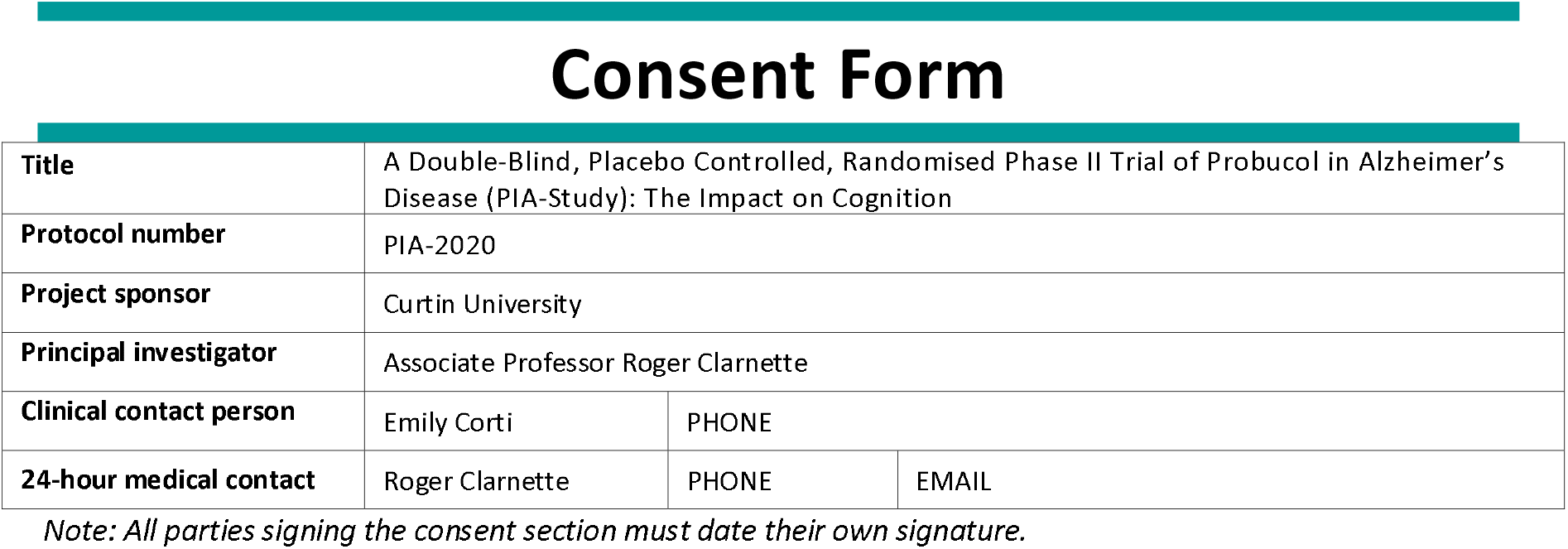

##### Declaration by participant

- I have read, or have had read to me, and I understand the General Information PICF as well as this Participant Information Sheet and Consent Form.
- I have had the opportunity to have a member of my family or another person present while the study is explained to me.
- I have had an opportunity to ask questions and I am satisfied with the answers I have received.
- I freely agree to participate in this study as described and understand that I am free to withdraw at any time during the study without affecting my future health care.
- I understand the purposes, procedures and risks of the research described in this Participant Information Sheet. Although I understand that the purpose of this study is to improve the quality of medical care, it has also been explained to me that my involvement may not be of any direct benefit to me.
- I understand that access may be required to my medical records for the purpose of this study as well as for quality assurance, auditing and in the event of a serious adverse event. I understand that such information will remain confidential.
- I understand that I will be randomised to either the Probucol (Lorelco™) or Placebo group, and that neither I nor the Study Doctor will know which group I am in.
- I understand and agree to my study partner providing information about my health to the Study Doctor.
- I consent to my local doctor being notified of my participation in this study and any clinically relevant information noted by the Study Doctor in the conduct of the study.
- I agree to adhere to the protocol requirements and restrictions as laid out in this Participant Information Sheet.
- I understand that I must use adequate contraception during the study. In the event of myself or my partner becoming pregnant, I must inform the Study Doctor immediately.
- I am 18 years of age and under 85 years of age.
- I understand that I will be given a signed copy of this document to keep.

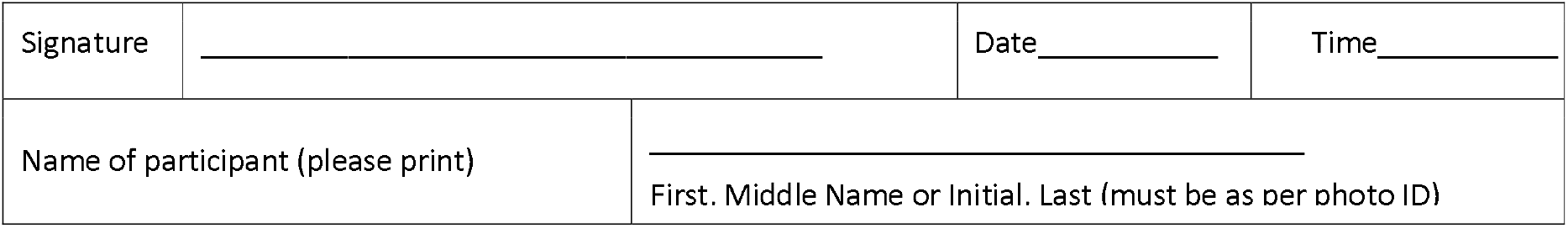

##### Declaration by trial doctor

I have given a verbal explanation of the clinical trial, its procedures and risks and I believe that the participant has understood that explanation.

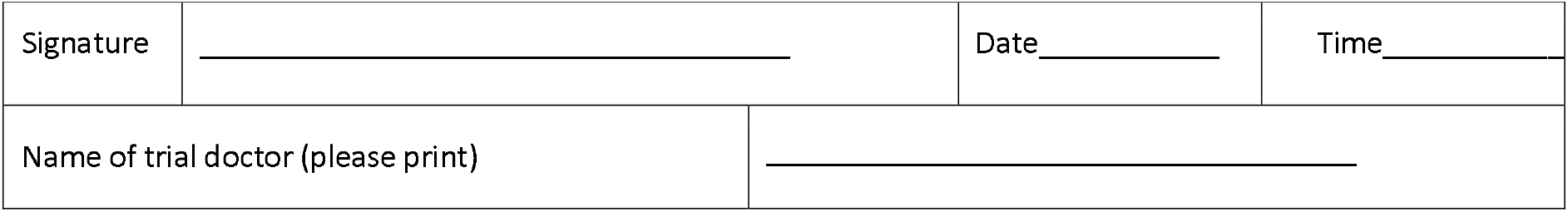

|  |  |  |  |
| --- | --- | --- | --- |
| Signature | _____ | Date_____ | Time_____ |
| Name of trial doctor (please print) | _____ |  |  |

#### Appendix B

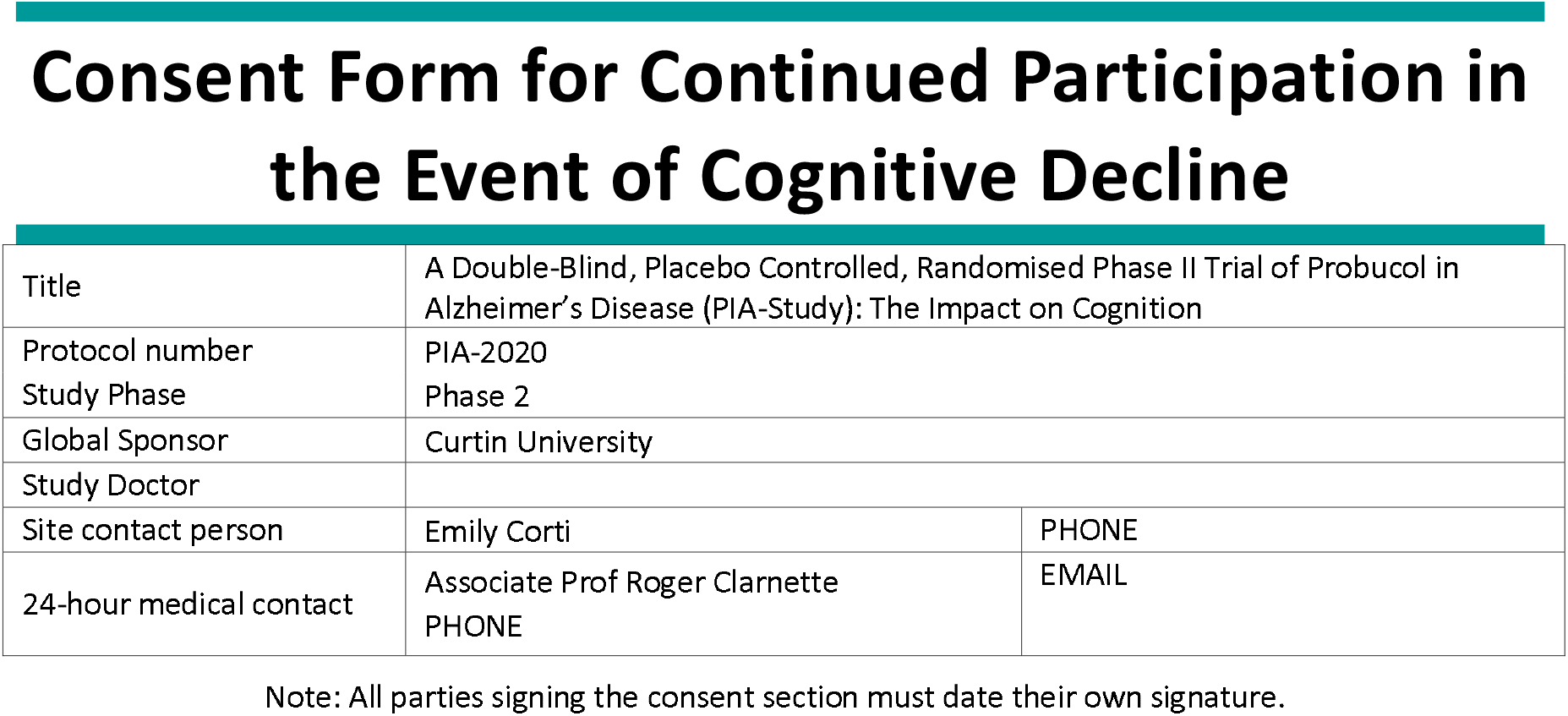

##### Declaration by Participant

- I have read, or have had read to me, and I understand the participant information and consent form.
- I have had the opportunity to have a member of my family or another person present while the study is explained to me.
- I have had an opportunity to ask questions and I am satisfied with the answers I have received.
- I freely agree to participate in this study as described and understand that I am free to withdraw at any time during the study without affecting my future health care.
- I understand that the Study Doctor has the right to withdraw me from the study at any point.
- I understand that I will be given a signed copy of this document to keep.

*I consent to continue my participation in the study in the event that my memory and thinking skills decline during the study*.

|  |  |  |  |
| --- | --- | --- | --- |
| Signature | _____ | Date_____ | Time_____ |
| Name of participant (please print) | _____<br>First Name, Middle Name or Initial, Last Name |  |  |

##### Declaration by Study Doctor

I have given a verbal explanation of the study, its procedures and risks and I believe that the participant has understood that explanation.

|  |  |  |  |
| --- | --- | --- | --- |
| Signature | _____ | Date_____ | Time_____ |
| Name of Study Doctor (please print) | _____ |  |  |

#### Appendix C

Consent Form for the Use and Storage of Blood Samples for Future Research
|  |  |  |
| --- | --- | --- |
| Title | A Double-Blind, Placebo Controlled, Randomised Phase II Trial of Probucol in Alzheimer's Disease (PIA-Study): The Impact on Cognition |  |
| Protocol number | PIA-2020 |  |
| Study Phase | Phase 2 |  |
| Global Sponsor | Curtin University |  |
| Study Doctor |  |  |
| Site contact person | Emily Corti | 0468 532 458 |
| 24-hour medical contact | Associate Prof Roger Clarnette<br>0415 956 611 |  |

Note: All parties signing the consent section must date their own signature.

##### Declaration by Participant

- I have read, or have had read to me, and I understand the participant information and consent form.
- I have had the opportunity to have a member of my family or another person present while the study is explained to me.
- I have had an opportunity to ask questions and I am satisfied with the answers I have received.
- I freely agree to participate in this study as described and understand that I am free to withdraw at any time during the study without affecting my future health care.
- I understand the purposes, procedures and risks of the research described in this Participant Information Sheet. Although I understand that the purpose of this study is to improve the quality of medical care, it has also been explained to me that my involvement may not be of any direct benefit to me.
- I consent to my local doctor being notified of my participation in this study and any clinically relevant information noted by the Study Doctor in the conduct of the study.
- I agree to adhere to the protocol requirements and restrictions as laid out in the main Participant Information Sheet and this Participant Information Sheet.
- I understand that I must use adequate contraception during the study. In the event of myself or my partner becoming pregnant, I must inform the Study Doctor immediately.
- I am over 18 years of age, or younger than 85 years of age.
- I understand that I will be given a signed copy of this document to keep.

*I consent to have additional blood samples taken for future research purposes in Alzheimer’s Disease and for blood samples taken from this study to be stored and used for future research purposes. I also consent to being contacted in the future about providing additional blood samples for future Alzheimer’s related projects*

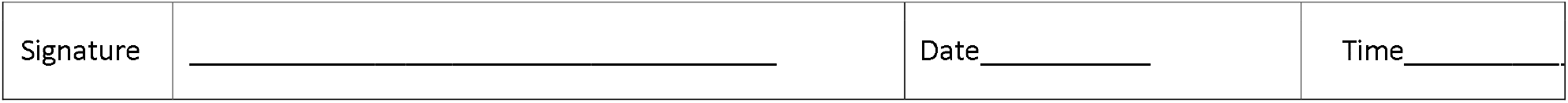
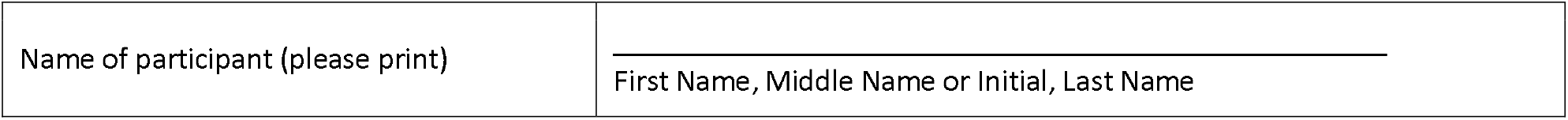

|  |  |  |  |
| --- | --- | --- | --- |
| Signature | _____ | Date_____ | Time_____ |

|  |  |
| --- | --- |
| Name of participant (please print) | _____ |
|  | First Name, Middle Name or Initial, Last Name |

##### Declaration by Study Doctor

I have given a verbal explanation of the study, its procedures and risks and I believe that the participant has understood that explanation.

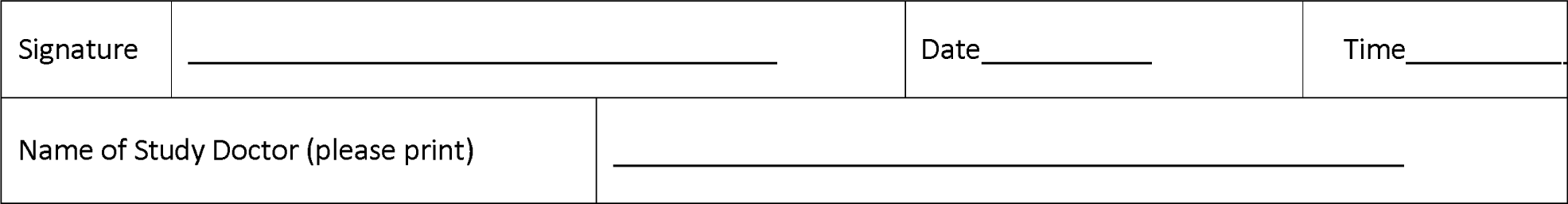

|  |  |  |  |
| --- | --- | --- | --- |
| Signature | _____ | Date_____ | Time_____ |
| Name of Study Doctor (please print) | _____ |  |  |

#### Appendix D

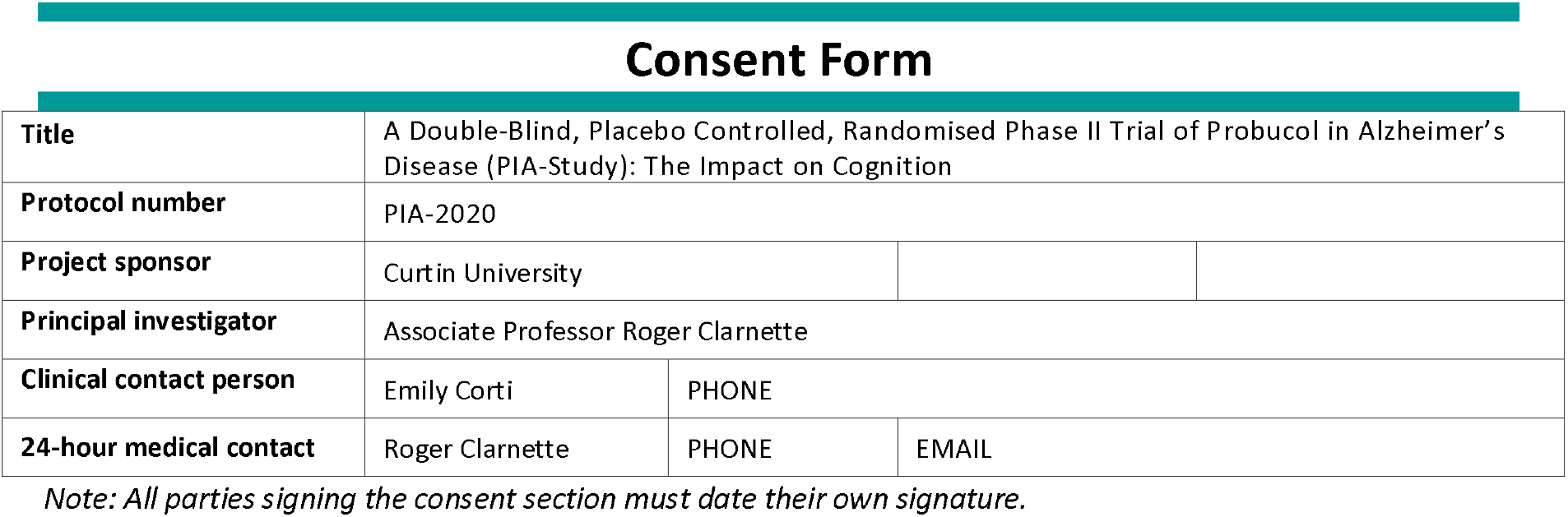

##### Declaration by study partner

- I have read, or have had read to me, and I understand the General Information PICF as well as this Partner Information Sheet and Consent Form.
- I agree to the above requirement of study partners, as set out in the Participant Information sheet and Consent form. I also acknowledge that at no time am I, as the study partner, receiving the treatment.
- I have had an opportunity to ask questions and I am satisfied with the answers I have received.
- I freely agree to participate in this study as described and understand that I am free to withdraw at any time during the study without affecting my future health care.
- I freely agree to meet the minimum study requirements which includes; attending one screening visit, completing a questionnaire at the start, at 6 months, and at the end of the study, and be available by phone monthly, as needed.
- I understand that by consenting to the minimum study requirements, the study participant may continue through the screening phase of the study and, if eligible, will receive the study medication.
- I understand that I will be given a signed copy of this document to keep.

|  |  |  |  |
| --- | --- | --- | --- |
| Signature | _____ | Date_____ | Time_____ |
| Name of study partner (please print) | _____ |  |  |
|  | First, Middle Name or Initial, Last (must be as per photo ID) |  |  |

##### Declaration by trial doctor

I have given a verbal explanation of the clinical trial, its procedures and risks and I believe that the study partner has understood that explanation.

|  |  |  |  |
| --- | --- | --- | --- |
| Signature | _____ | Date _____ | Time _____ |
| Name of trial doctor (please print) |  | _____ |  |

## Notes

### Competing Interest Statement

The authors have declared no competing interest.

### Clinical Trial

ACTRN12621000726853

### Author Declarations

Approved by BellBerry Human Research Ethics Committee. The approval number is 2019-11-1063

